# Two to Tango: Are Spouses’ Perception of Women’s Empowerment Associated with Concordance on Fertility Desires?

**DOI:** 10.64898/2026.07.22.26358718

**Authors:** Bhavita Kumari

## Abstract

**Introduction:** The current study examined the association between husbands’ and wives’ perceptions of women’s empowerment, focusing on household decision-making, attitudes toward IPV, and fertility desires in Pakistan.

**Methods:** Data from currently married couples participating in the Pakistan DHS (2017–18) were included in the analysis (n=3,027). The level of concordance on fertility desires was assessed using Kappa Coefficient. Principal component analysis (PCA) was applied to assess the internal consistency and dimensionality of household decision-making and IPV attitude items; items within each construct were subsequently summed to generate categorical empowerment scores used in the regression analyses. Relative risk ratios were estimated for perceived empowerment on spousal concordance on fertility desires using multinomial logistic regressions.

**Results:** The study found a moderate level of spousal agreement on fertility desires (Kappa=0.51). Among significant findings: wife’s perception of her own high decision-making authority was associated with more than a two-fold increase in wife-only discordance (wife wanting more children, husband wanting no more), compared to both spouses wanting more children.

**Conclusion:** The study contributes to the understanding of couple’s gendered dynamics and how they may influence fertility desires. Such knowledge informs efforts to increase both spouses’ involvement in reproductive decision-making and behaviors.

## Introduction

Traditionally, family planning research in Pakistan has focused primarily on women’s perspectives, often overlooking men’s and couples’ viewpoints (1,2). Studies on couple dynamics in reproductive decision-making reveal significant disagreements, particularly regarding contraceptive use and family size preferences (3,4). Fertility decisions typically involve joint decision-making, yet large-scale surveys often rely on women’s reports of their partners’ fertility desires, which may not accurately reflect their true preferences (5). Recognizing the dyadic nature of fertility decisions, incorporating both spouses’ desires can improve the understanding of reproductive behaviors (6). Spousal concordance on fertility desires has been linked to higher contraceptive use (7), while discordance can arise due to gendered social norms, power dynamics, and social desirability bias (8). Men’s perspectives are crucial in shaping reproductive outcomes, as male-dominated decision-making often prioritizes husbands’ desires over that of wives’ (5). Engaging discordant couples through spousal communication interventions may help align fertility desires and enhance contraceptive use (7).

In Pakistan, fertility rates have declined from around seven births per woman in 1990 to 3.6 in 2017-18, but progress has stalled in recent years, with modern contraceptive prevalence rates stagnant at 25% (9). Pronatalist norms persist in the Pakistani patriarchal society (10). Despite this, social changes over the past two decades, including increased female literacy (from 21% in 1990 to 47% in 2011-12) and labor force participation (rising to 24.3% in 2011-12), have empowered women, potentially influencing fertility preferences (11). However, these societal shifts have also introduced tensions in spousal fertility desires, especially when women’s increasing autonomy leads to smaller ideal family sizes, while male-dominated norms still favor larger families (12).

While spousal concordance on fertility desires has been linked to contraceptive use (13), limited research has explored the predictors of discordance. Studies suggest that higher education and economic opportunities enhance women’s bargaining power (5), yet spousal discordance remains influenced by patriarchal norms and power imbalances (14). Evidence on the relationship between women’s empowerment and fertility discordance is mixed: while some studies report no significant association, others find that education is associated with greater spousal alignment on family size preferences. Reproductive control and intimate partner violence have been shown to limit women’s negotiating power over contraception, reinforcing male-dominant fertility preferences (5). Given the lack of couple-level data, further research is needed to understand how spousal perceptions of women’s empowerment affect fertility preferences, contraceptive use, and reproductive decision-making in Pakistan. This study aims to bridge this gap by examining the extent and determinants of spousal discordance on fertility desires. Specifically, the objectives of the study include: 1) assess the level of spousal concordance on fertility desires among currently married couples in Pakistan, and 2) explore the association between spouses’ perceptions of women’s empowerment and spousal concordance/discordance on fertility desires among currently married couples in Pakistan through two hypotheses: 2.1) *<u>Women’s own perception of higher empowerment is associated with increased wife-only discordance on fertility desires (wife wanting more children, husband wanting no more) among currently married couples in Pakistan,</u>* and<u> (2.2.) *Husbands’ perception of women’s empowerment moderates the association between women’s own perception of their empowerment and spousal concordance on fertility intentions, whereby both spouses desired no more children*</u>

### Conceptual Framework

This study is informed by Connell’s Theory of Gender and Power, as adapted by Wingood and DiClemente, due to its comprehensive integration of gender dynamics across multiple levels and its impact on women’s health outcomes (15). The framework incorporates various aspects of women’s empowerment within the context of reproductive behaviors. The theory has been widely applied in research exploring the connection between gender inequality and reproductive behaviors in the developing world (16,17). It outlines three key social structures that shape gender relations: the sexual division of labor, the sexual division of power, and the structure of cathexis (15). Figure 1 shows determinants of spousal concordance on fertility desires and their relationship with each other. Note: Figure 1 presents the broader theoretical framework as conceptualized by Wingood and DiClemente (15). Several constructs depicted therein — including polygamy, ethnic minority status, religious affiliation, and spousal communication on reproductive decisions — were not available in the PDHS 2017-18 data and are therefore not operationalized in the present analysis. The constructs empirically examined are described in the Methods section (Table 2).

**Figure 1.**
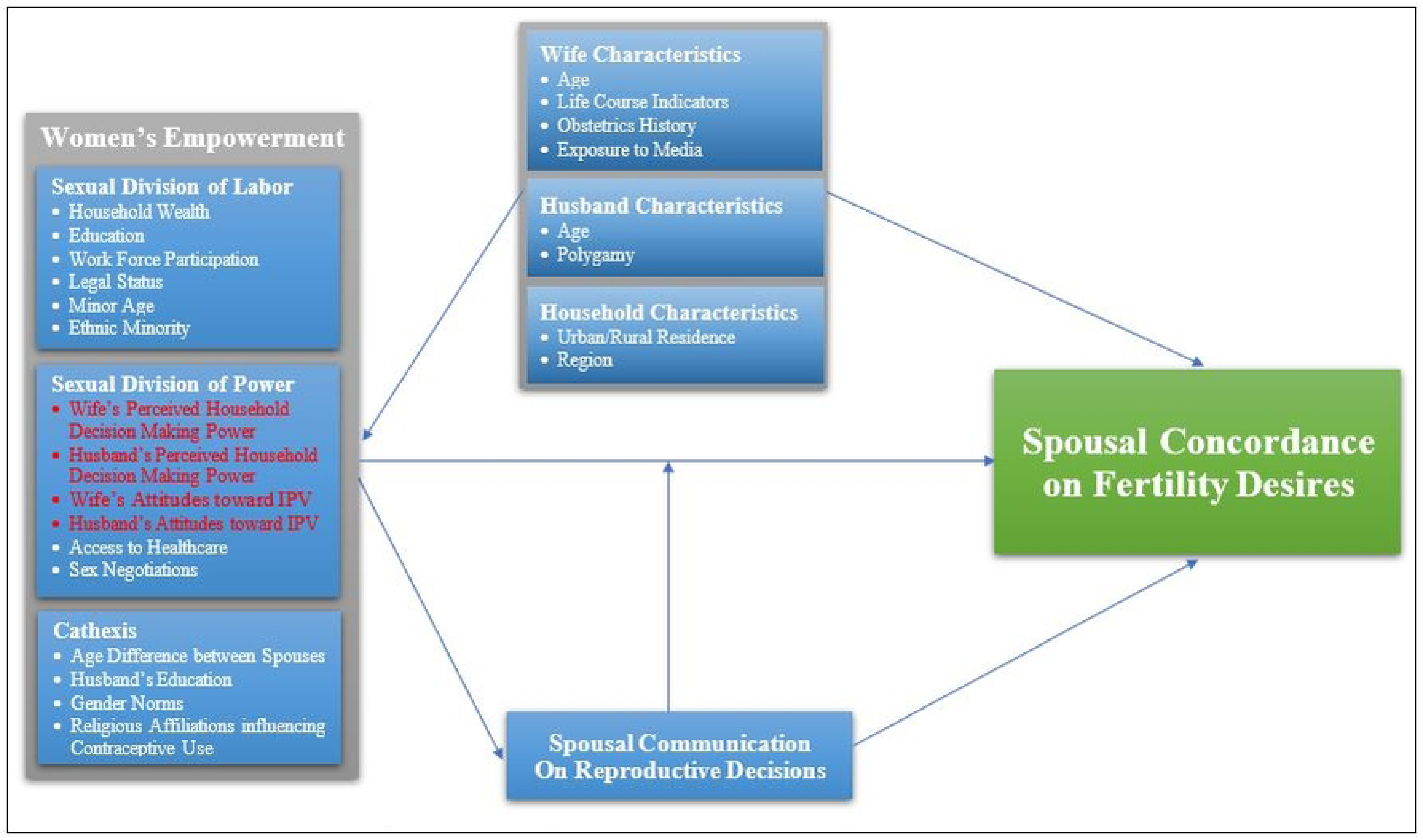
Conceptual framework of relationship between women’s empowerment reported by spouses and couple factors and spousal concordance on fertility desires in Pakistan.

The ‘sexual division of power’, that is imbalance of decision-making control and inequities power, highlights an imbalanced spousal power dynamic. Women’s perceived household decision-making power is positively associated with enhanced spousal communication, potentially improving spousal concordance on reproductive desires and behaviors (18). When husbands perceive themselves as decision-makers, wife’s healthcare utilization is likely to reduce (19). Economically disadvantaged women are at increased risk of IPV. They are more likely to feel apprehensive discussing reproductive choices and be submissive to their husband’s fertility desires (20,21). Hence, investigating attitudes of both spouses is essential for understanding couple’s fertility desires.

The second empowerment structure, *sexual division of labor*, refers to differences in economic opportunities between men and women. It comprises factors that contribute to the educational and financial empowerment of women. The literature suggests that fertility desires are affected by sexual division of labor, which includes household wealth, education, age, ethnicity and employment, and legal status (15,22). However, reproductive choices are also affected by husband’s education(5), hence the need to explore the spousal difference of labor. The third division of women’s empowerment, *cathexis,* refers to the emotional and relational dimensions of power within intimate partnerships. In this study, it is operationalized using the spousal age difference and husband’s education, following prior applications of the theory in LMIC contexts (23). Husband’s education shapes his attitudes toward gender norms, power-sharing, and communication within the household, and has been shown to influence women’s reproductive autonomy (5), justifying its classification under cathexis rather than the sexual division of labor. (23).

## Methods

This study was conducted using the couple data from the PDHS 2017-18. The approval to use the anonymous dataset was authorized on 22^nd^ January, 2018. The survey sample included currently married women aged 15-49 from selected households and their husbands. Couples where either of the spouses reported being sterilized or infecund are excluded. Fertility desires were measured by responses of husband and wife to a question about desires for more children as described in Table 1. In the DHS, husband, and wife were asked separately, “would you like to have (a/another child, or would you prefer not to have any (more) children?” Spousal concordance on fertility desire were categorized into four groups as follows: 1) “concordant (want more)” – both spouses desired more children; 2) “husband+ discordant” – the husband wanted more children, but the wife did not; 3) “wife+ discordant” – the wife wanted more children, but the husband did not; 4) “concordant (no more)” – both spouses wanted to stop having children.

**Table 1:** Cross-tabulation showing categories and level of spousal concordance on fertility desires, as measured in PDHS 2017-18. (N = 3,027)

|  |  | Husband |  | Total % (n) | p-value |
| --- | --- | --- | --- | --- | --- |
|  |  | *Wants More % (n)* | *Wants No More % (n)* |  |  |
| Wife | *Wants More* | Concordance (Want More)<br>54.3 (1,645) | Wife+ Discordance<br>8.6 (259) | 62.4 (1,904) | <0.01 |
|  | *Wants No More* | Husband+ Discordance<br>13.8 (418) | Concordance (Want No More)<br>23.3 (705) | 37.6 (1,123) |  |
| Total |  | 68.2 (2,063) | 31.8 (964) | 100 (3,027) |  |

| Agreement | Expected Agreement | Kappa Value | p-value |
| --- | --- | --- | --- |
| 77.6% | 54.7% | 0.51 | <0.01 |

p-value based on chi-square test PDHS Couple Survey 2017-18 (not weighted)

Key independent variables (household decision-making and attitudes toward IPV as reported by both spouses) and other domains of women’s empowerment as identified based on the Theory of Gender and Power are described in Table 2. Empowerment variables were recoded to dichotomous scales such that a higher score reflects a more empowered woman. Responses to household decision-making questions were recoded as 0 = husband/someone else deciding and 1=wife deciding alone/jointly. Similarly, responses for attitudes toward IPV were recoded as 0=justified and 1=not justified. Responses for access to healthcare are recoded as 0=big problem and 1= not a big problem. Responses for sex negotiations were recoded as 0=cannot negotiate, and 1=can negotiate. Legal status was recoded as 0=no ownership and 1=ownership of land, house, or both. The individual questions under each concept are listed in Table 2.

**Table 2:**
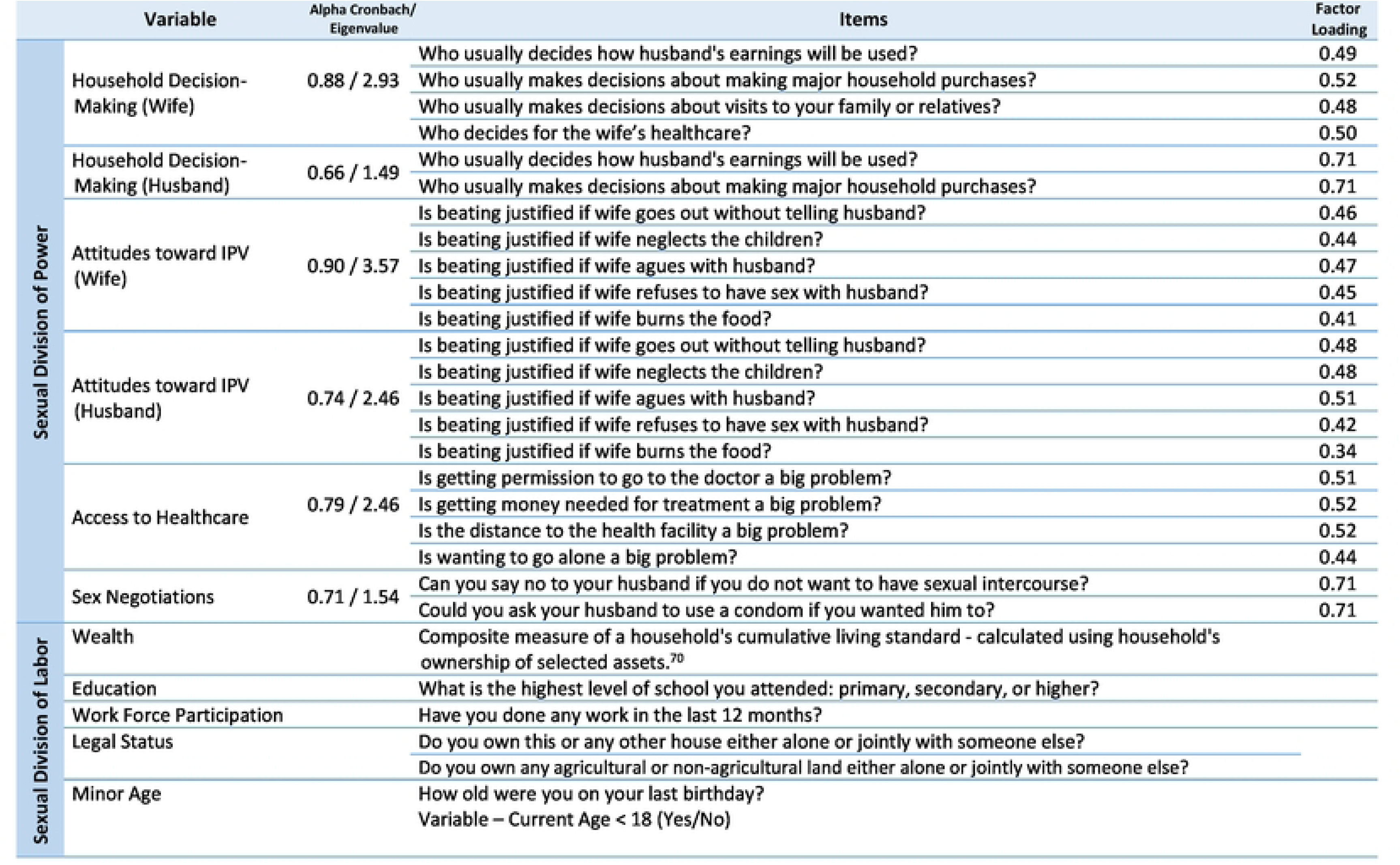
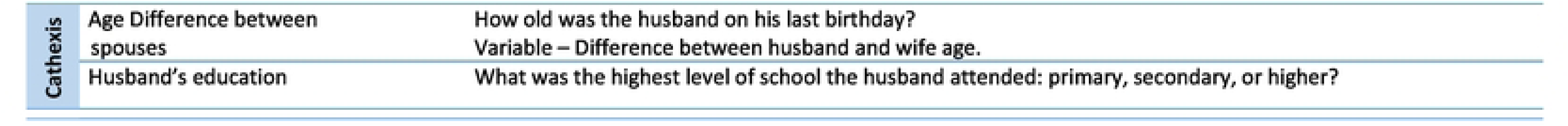
Description of the empowerment variables, according to the Theory of Gender and Power, and principal component analysis of Items, as measured in Pakistan DHS 2017-18 and reported by both spouses. (n = 3,027)

|  | Variable | Alpha Cronbach/<br>Eigenvalue | Items | Factor<br>Loading |
| --- | --- | --- | --- | --- |
| Sexual Division of Power | Household Decision-Making (Wife) | 0.88 / 2.93 | Who usually decides how husband's earnings will be used? | 0.49 |
|  |  |  | Who usually makes decisions about making major household purchases? | 0.52 |
|  |  |  | Who usually makes decisions about visits to your family or relatives? | 0.48 |
|  |  |  | Who decides for the wife's healthcare? | 0.50 |
|  | Household Decision-Making (Husband) | 0.66 / 1.49 | Who usually decides how husband's earnings will be used? | 0.71 |
|  |  |  | Who usually makes decisions about making major household purchases? | 0.71 |
|  | Attitudes toward IPV (Wife) | 0.90 / 3.57 | Is beating justified if wife goes out without telling husband? | 0.46 |
|  |  |  | Is beating justified if wife neglects the children? | 0.44 |
|  |  |  | Is beating justified if wife argues with husband? | 0.47 |
|  |  |  | Is beating justified if wife refuses to have sex with husband? | 0.45 |
|  |  |  | Is beating justified if wife burns the food? | 0.41 |
|  | Attitudes toward IPV (Husband) | 0.74 / 2.46 | Is beating justified if wife goes out without telling husband? | 0.48 |
|  |  |  | Is beating justified if wife neglects the children? | 0.48 |
|  |  |  | Is beating justified if wife argues with husband? | 0.51 |
|  |  |  | Is beating justified if wife refuses to have sex with husband? | 0.42 |
|  |  |  | Is beating justified if wife burns the food? | 0.34 |
|  | Access to Healthcare | 0.79 / 2.46 | Is getting permission to go to the doctor a big problem? | 0.51 |
|  |  |  | Is getting money needed for treatment a big problem? | 0.52 |
|  |  |  | Is the distance to the health facility a big problem? | 0.52 |
|  |  |  | Is wanting to go alone a big problem? | 0.44 |
|  | Sex Negotiations | 0.71 / 1.54 | Can you say no to your husband if you do not want to have sexual intercourse? | 0.71 |
|  |  |  | Could you ask your husband to use a condom if you wanted him to? | 0.71 |
| Sexual Division of Labor | Wealth |  | Composite measure of a household's cumulative living standard - calculated using household's ownership of selected assets.<sup>70</sup> |  |
|  | Education |  | What is the highest level of school you attended: primary, secondary, or higher? |  |
|  | Work Force Participation |  | Have you done any work in the last 12 months? |  |
|  | Legal Status |  | Do you own this or any other house either alone or jointly with someone else? |  |
|  |  |  | Do you own any agricultural or non-agricultural land either alone or jointly with someone else? |  |
|  | Minor Age |  | How old were you on your last birthday?<br>Variable – Current Age < 18 (Yes/No) |  |

|  |  |  |
| --- | --- | --- |
| Cathexis | Age Difference between spouses | How old was the husband on his last birthday?<br>Variable – Difference between husband and wife age. |
|  | Husband's education | What was the highest level of school the husband attended: primary, secondary, or higher? |

The individual measures of women’s empowerment were used to construct latent measures of decision-making powers perceptions and attitudes toward IPV of husband and wife and other domains of empowerment, using principal component analysis (as shown in Table 2). Internal consistency of each construct was assessed by Cronbach’s alpha; an alpha value greater than the cutoff of 0.7 and Principal Component Analysis (PCA) eigenvalue greater than 1.0 were considered high internal consistency. Table 2 reports the Cronbach’s alpha and eigenvalues for the given constructs, indicating high internal consistency among the items for these constructs. Factor loadings greater than 0.40 were used to determine the match between the items and each construct.

Based on the PCA results, these individual measures were summed to generate a score ranging from 0 to 4. The scores were then recoded into categorical variables: Household decision-making variables, reported by both wife and husband, were recoded as 0= no decision-making (women not having any say in any situations), 1=moderate decision-making (women having a say in 1-3 situations), and 2=high decision-making (women having a say in all 4 situations). Attitudes toward IPV, reported by both wife and husband, were recoded as 0=no intolerance (women’s report of “not justified” in 0-1 situation), 1=moderate intolerance (beating “not justified” in 2-4 situations), and 2=high intolerance (beating “not justified” in 5 situations). Access to healthcare was recoded as 0=big problem (4 problems in accessing healthcare), 1=moderate problem (2-3 problems in accessing healthcare), and 2=no problem (1 or no problem in accessing healthcare) and sex negotiations as 0=cannot negotiate and 1= can negotiate (in one or both situations).

Using Stata 12, we used the *-svy* set of commands to account for the complex two-stage cluster sampling design of the PDHS. For research objective 1, the level of the spousal concordance on fertility desires is analyzed using Kappa statistic. The strength of agreement was categorized following Landis and Koch’s criterion (24). Correlation analyses were conducted to identify potentially correlated variables. More than 0.60 was considered to be high correlation among variables, and wealth, husband age, total children born, and age at first birth were removed from the analysis. There was a risk of collinearity between the husband’s and wife’s perceptions of women’s empowerment; however, we found the collinearity coefficient between them to be low (0.25). Similarly, there was moderate collinearity between husband and wife’s fertility desires (0.51).

Chi-square tests of independence for categorical variables and one-way ANOVA were used for continuous variables for bivariate analyses. For research objective 2, multinomial logistic regressions were performed to assess the association between spousal concordance on fertility desires and i) women’s self-perceived empowerment for hypothesis 2.1, and ii) interactions of both spouses’ perceptions of household decision-making reports for hypothesis

### 2.2. Relative risk ratios were reported for both analyses

#### Women’s Empowerment and Spousal Concordance on Fertility Intentions

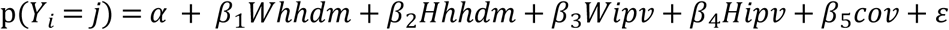

#### Interaction Effect of Husband’s and Wife’s Perception of Women’s Empowerment and Spousal Concordance on Fertility Intentions

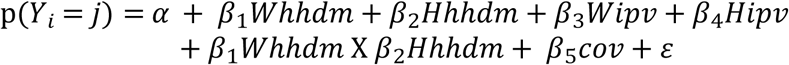

Where: 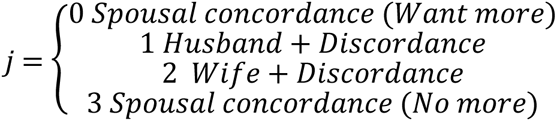

Whhdm = Categorical empowerment score for household decision-making as reported by the wife (derived from PCA-validated summed items)
Hhhdm = Categorical empowerment score for household decision-making as reported by the husband (derived from PCA-validated summed items)
Wipv = Categorical empowerment score for wife’s attitude toward intimate partner violence (derived from PCA-validated summed items)
Hipv = Categorical empowerment score for husband’s attitude toward intimate partner violence (derived from PCA-validated summed items)
Cov = Vector of individual, spousal, and household covariates

## Results

Frequencies and percentage of spousal concordance and discordance on fertility desires are given in Table 1. The Kappa value for agreement between husband and wife’s report of fertility desires was 0.51, suggesting a moderate level of spousal agreement in this sample. Descriptive statistics and bivariate analysis for the study sample, including the individual characteristics of the husbands and wives and their household characteristics, are shown in Table 3.

**Table 3:** Description of socio-demographic variables potentially associated with women’s employment, as measured in PDHS 2017-18 (N = 3,027)

| Variables | Categories | Wife % (n) | Husband % (n) |
| --- | --- | --- | --- |
| Age | *15-19* | 6.0 (187) | 1.4 (44) |
|  | *20-24* | 20.8 (569) | 9.5 (264) |
|  | *25-29* | 23.8 (760) | 20.2 (571) |
|  | *30-34* | 22.2 (634) | 20.8 (670) |
|  | *35-39* | 16.5 (527) | 20.8 (619) |
|  | *40-44* | 6.9 (248) | 14.1 (436) |
|  | *45-49* | 3.9 (102) | 13.2 (423) |
| Education | *No Education* | 46.8 (1,498) | 25.3 (720) |
|  | *Primary* | 14.9 (416) | 20.4 (512) |
|  | *Secondary or* | 38.2 (1,113) | 54.4 (1,795) |
| Life Course Indicators |  | *1st: 27.8* | *No: 97.0 (2,958)* |
|  |  | *2nd/3rd: 72.2* | *Yes: 3.0 (69)* |
| *Exposure to Media* | *No* | 74.5 (2369) | 50.8 (1,727) |
|  | *Yes* | 25.5 (658) | 49.2 (1,300) |
| **Continuous Variables** |  | **Mean ± SD** |  |
| Life Course Indicators | Age at Marriage | 19.5 ± 4.1 |  |
|  | Age at First Birth | 21.2 ± 4.1 |  |
| Obstetric History | Number of children ever born | 3.1 ± 2.4 |  |
|  | Number of Living sons | 1.4 ± 1.3 |  |
| **Household Characteristics** |  | **Household % (n)** |  |
| Wealth Index | *Poorest* | 18.1 (570) |  |
|  | *Poorer* | 19.5 (684) |  |
|  | *Middle* | 19.3 (572) |  |
|  | *Richer* | 21.1 (579) |  |
|  | *Richest* | 21.9 (622) |  |
| Residence | *Urban* | 40.2 (1,522) |  |
|  | *Rural* | 59.8 (1,505) |  |

Tabulations account for survey weights

Table 4 presents results of multinomial logistic regressions examining associations between women’s empowerment variables and spousal concordance on fertility desires, unadjusted (Model I) and adjusted for covariates (Model II). Of 27 adjusted coefficients, 23 were not statistically significant. Among significant associations after adjustment: when wife perceived high decision-making authority, wife-only discordance (wife wanting more children) was 2.06 times more likely than both spouses wanting more (RRR=2.06, 95% CI: 1.20–3.55; p<0.01). When the husband perceived his wife’s decision-making as high, wife-only discordance was 62% less likely (RRR=0.38, 95% CI: 0.21–0.66; p<0.01). High husband intolerance toward IPV increased husband-only discordance 2.48-fold (RRR=2.48, 95% CI: 1.08–5.66; p<0.05). Wife’s ability to negotiate sex increased concordance on wanting no more children 2.09-fold (RRR=2.09, 95% CI: 1.40–3.13; p<0.01).

**Table 4:**
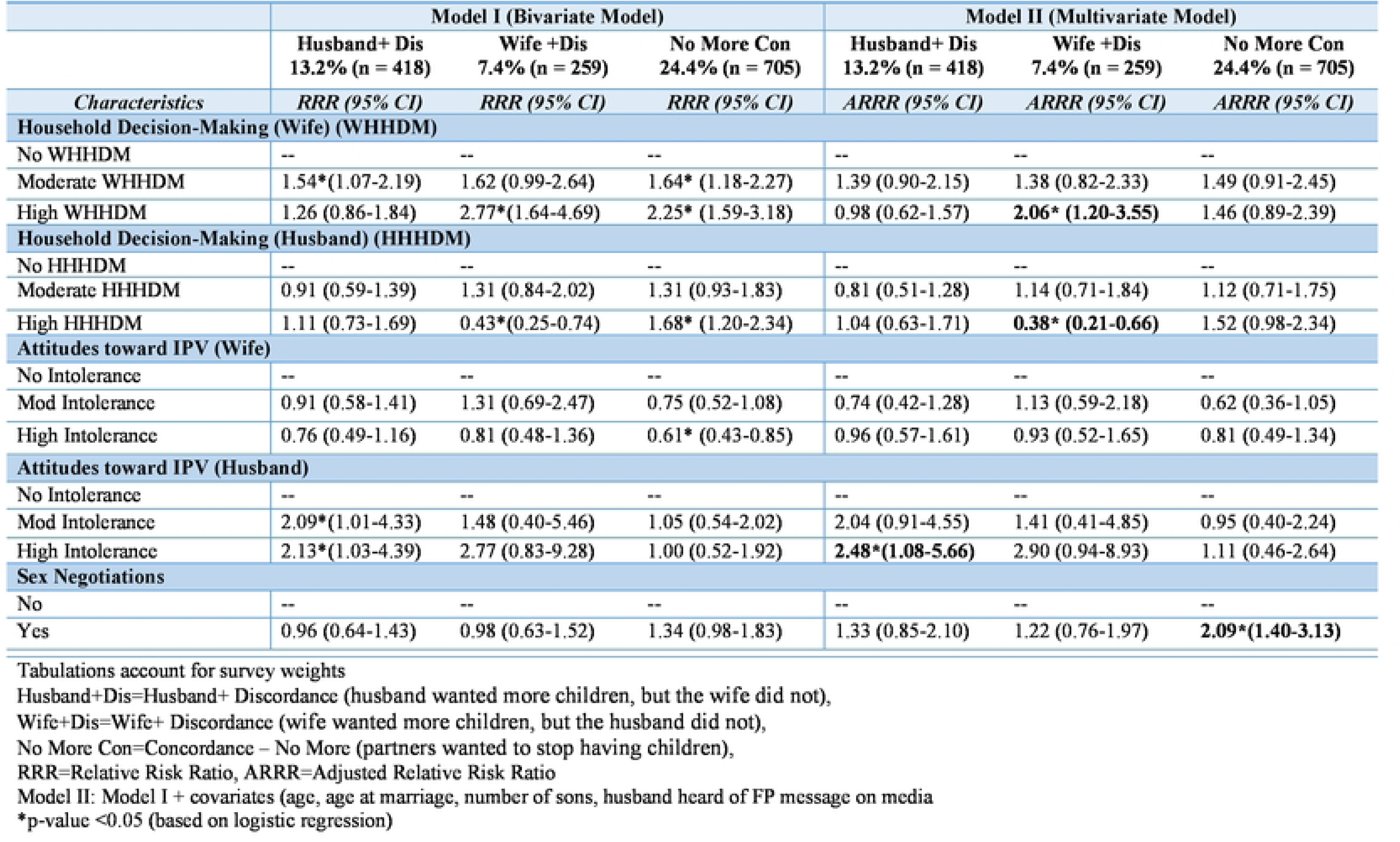
Multinomial logistic regression results for the association between empowerment characteristics (reported by both spouses) and spousal concordance on fertility desires, PDHS 2017-18 (N=3,027) (Reference category: concordance on wanting more children)

Figure 2 shows the results from the interaction analysis between both spouses’ reports of wife’s empowerment. When both husband and wife perceived wife’s moderate decision-making authority, the likelihood of wife only wanting more children was 79% lower (RRR=0.21, 95% CI: 0.07-0.61; p<0.01), and that neither spouse wanting more children was 68% lower (RRR=0.32, 95% CI: 0.11-0.94; p<0.05)., compared to both spouses wanting more children. Similarly, when the wife perceived high decision-making authority for herself & the husband perceived moderate decision-making authority, the likelihood of neither spouse wanting more children was 77% lower than the likelihood of both spouses wanting more children (RRR=0.23, 95% CI: 0.09-0.59; p<0.01). When both husband and wife perceived high decision-making authority for the woman, the likelihood of neither spouse wanting more children was 78% lower than the likelihood of both spouses wanting more children (RRR=0.22, 95%CI: 0.08-0.61; p<0.01).

**Figure 2:**
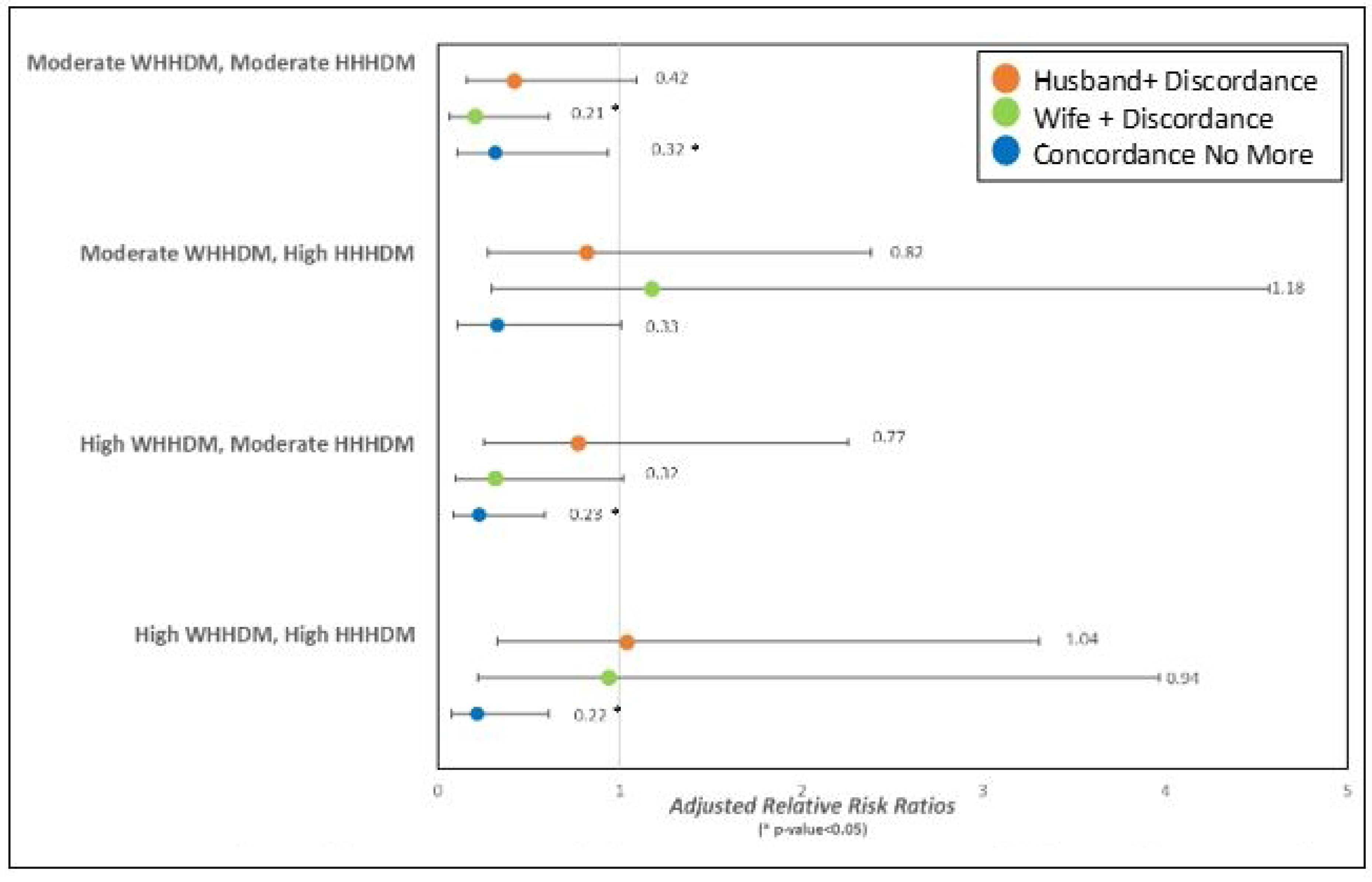
Multinomial logistic regression results for association between interaction of both spouses’ perceptions of household decision-making (HHDM) and spousal concordance on fertility intentions, Pakistan DHS 2017-18 (N=l4,502) WHHDM (Wife Household Decision-Making). HHHDM (Husband Household Decision-Making)

## Discussion

This study aimed to examine the level of spousal concordance on fertility desires and its associations with perceptions of women’s empowerment from the perspectives of both spouses among currently married couples in Pakistan. The study suggests a moderate level of concordance between spouses on fertility desires, Kappa value being 0.51, underlining the importance of collecting information on fertility desires from both spouses. The level of spousal agreement varied greatly across other LMICs, including India and Sub-Saharan countries; Kappa values ranging from as low as 0.37 in Ethiopia (25) to as high as 0.75 in India (13). However, most of the studies reporting Kappa values were conducted in specific towns or villages with relatively small sample sizes, compared to our nationally representative, large sample. A central finding requiring explicit acknowledgment is that 23 of the 27 adjusted regression coefficients in Table 4 were not statistically significant, meaning the empowerment constructs examined largely failed to predict variation in spousal concordance outcomes in this sample. This null pattern likely reflects several factors. First, in settings with strong pronatalist norms, fertility preferences may be shaped primarily by shared social expectations rather than within-couple power dynamics, producing similar fertility desires regardless of empowerment level. Second, the PDHS instruments were not designed to capture the full complexity of reproductive autonomy; key determinants such as spousal communication quality, relationship satisfaction, and community-level gender norms are absent from the data. Third, the categorical operationalization of empowerment scores using item-count thresholds rather than continuous latent variables may reduce statistical sensitivity. These findings underscore the need for studies using purpose-built instruments and longitudinal or qualitative designs to adequately test empowerment-concordance relationships.

The study found that wife’s self-report of high decision-making authority was associated with an increase by more than two folds in discordance, whereby the wife wanted more children, and the husband wanted no more, compared to couples, whereby both spouses wanted more children. On the other hand, the husband’s perception of wife’s high decision-making authority was associated with around 60% decrease in discordance, whereby the wife wanted more children, and the husband wanted no more, compared to couples, whereby both spouses wanted more children. From the interaction analysis (Figure 2), when both spouses perceived wife’s moderate decision-making authority, the likelihood of neither spouse wanting more children was approximately 70% lower (RRR=0.32, 95% CI: 0.11–0.94); when both perceived high decision-making authority, this was approximately 80% lower (RRR=0.22, 95% CI: 0.08–0.61), relative to concordance whereby both spouses wanted more children.

In patriarchal societies, concordance on fertility desires is high as women are often ‘taught’ since childhood to accept their husband’s preferences and often not encouraged to voice their opinions (26). The societal gendered norms obstruct spousal communication on reproductive decisions, leading to “concordance” on fertility intentions. Furthermore, women’s respect in the society is often proportional to the number of children she has given birth to, especially sons (26), leading to a higher fertility intention among women. On the other hand, generally husbands pave the path to break any gender norms in the household, leading to increased spousal communication. This change in the household dynamics encourages women to express their opinions and choices freely (10).

Literature suggests that women’s education was associated with economic autonomy and decision-making authority at the household level (27), but that did not necessarily translate automatically into reproductive autonomy (28) The association of education and informed reproductive choices is influenced by gendered norms in a patriarchal society where men’s authority was paramount (10). Women may be more likely to compromise on their fertility desires to avoid conflict with their spouses or adhere to societal norms (28). Education provides women with agency and resources to make informed decisions about their reproductive behaviors (14).

Pakistan is undergoing a gradual gender transition, with women’s education and employment rates slowly improving (29). Even though gender roles are changing at the societal level in Pakistan, the dynamics at the household level remain gendered (30). Changes in societal norms seem to be transiting very slowly at the couple level, probably creating more spousal drift at this initial stage of transition. Hence, it remains ambiguous if women’s empowerment improves reproductive autonomy. Our findings suggest that even when women’s empowerment is considered from both spouses’ perspectives, it is crucial to consider spousal communication and gender norms in society.

This study provides nationally representative evidence on the dyadic nature of fertility desires in Pakistan, demonstrating a moderate level of spousal concordance and identifying household decision-making authority as the most consistently associated empowerment dimension. The finding that most empowerment constructs did not significantly predict concordance underscores the need for couple-level family planning interventions that address shared social norms and spousal communication alongside individual empowerment. Fertility desires are a precursor for reproductive behaviors; level of concordance on couples’ fertility desires can help predict future fertility trends (31). Hence, this information can help predict the demand for family planning and provide information on fertility trends. The moderate level of concordance on fertility desires indicates the importance of bringing the focus to couple-level interventions. The study informs policies to incorporate components on women’s empowerment, especially household-level power dynamics. It will enable the women to negotiate reproductive intentions with their husbands and subsequently empower women towards reproductive autonomy. There is a need to train men to make space for empowering women while encouraging men’s inclusion in family planning programs focusing on couple counseling and encouraging spousal communication about reproductive decisions.

The representative nature of the survey is the key strength, allowing generalization of results to the married couples of reproductive ages in Pakistan. There have been very few studies in Pakistan which report the interaction of the fertility desires of both spouses. This is one of the first studies that explored the associations between empowerment constructs and spousal concordance on fertility desires. The PDHS 2017-18 collected information on most of the empowerment factors as mentioned in the conceptual framework based on the Theory of Gender and Power, studying women’s empowerment as a holistic construct and from both spouses’ perspectives.

One of the limitations of PDHS surveys is lack of information on spousal communication on fertility desires and reproductive behaviors as well as societal-level characteristics, like gender norms and religious beliefs. Spousal communication on reproductive choices and decisions is expected to improve concordance on fertility intentions and contraceptive use.. It is crucial to explore how gendered norms at the societal level translate into reproductive autonomy at the couple level. In addition, the PDHS does not collect information on quantifying the couple’s fertility desires, that is, how many more children the spouses want. Finally, qualitative research should seek to explain further the perceptions of husband and wife regarding women’s empowerment and the processes of how empowerment at all levels affects their agreement regarding fertility desires.

Reporting issues like response bias due to social desirability can lead participants to underreport their actual desire for more children.(32,33) There can be ‘hidden discordance’ within concordance; for example, couples may want more children, but there could be disagreement over the exact number or timing. Social desirability can lead to underestimating the fertility desires in either spouse, usually the wife; it can lead to discordance, whereby the husband wants more children.

## Conclusion

The study contributes to understanding couples’ gender dynamics and how they may influence fertility desires, especially in a society with dynamic gendered roles. Such understanding informs efforts to increase both spouses’ involvement in reproductive practices. Future research should employ Actor-Partner Interdependence Models (APIM) using dyadic data to disentangle each spouse’s independent contribution to fertility concordance, and should incorporate spousal communication quality and community-level gender norm measures currently absent from large-scale surveys such as the PDHS.

## Data Availability

DHS Datasets were used for the analysis which are available as public datasets.

## Ethical Statement

Not applicable

## Disclaimer

No disclaimer

